# National Estimates for the Percentage of all Readmissions With Demographic Features, Morbidity, Overall and Gender-specific Mortality of Transcutaneous vs Open Surgical Tricuspid Valve Replacement/Repair

**DOI:** 10.1101/2023.04.25.23289124

**Authors:** Muhammad Shayan Khan, Abdul Baqi, Ghulam Mujtaba Ghumman, Waqas Ullah, Jay Shah, Yasar Sattar, Tanveer Mir, Zain Sheikh, Ayesha Tahir, Fnu Salman, Moaaz Baghal, Kritika Luthra, Vinod Khatri, Zainulabedin Waqar, Mohammed Taleb, Syed Sohail Ali

**Affiliations:** Department of Cardiology, Mercy Saint Vincent Medical Center, Toledo, OH; Department of Internal Medicine, Mercy Saint Vincent Medical Center, Toledo, OH; Department of Cardiology, Thomas Jefferson University Hospitals, Philadelphia, PA; Department of Cardiology, West Virginia University, Morgantown, WV; Department of Cardiology, Detroit Medical Center/Wayne State University, Detroit, MI; Department of Internal Medicine, Franciscan Health Care, Michigan, Indiana; Department of Pulmonology, Mercy Saint Vincent Medical Center, Toledo, OH

**Author notes:** **Corresponding Author:** Muhammad Shayan Khan, MD, Address: 2213 Cherry Street, Toledo, OH, 43608. **Funding:** This research did not receive any specific grant from funding agencies in the public, commercial, or not-for-profit sectors. **Declarations of Interest:** None.

## Abstract

**Aims:** To determine national estimates for the percentage of all readmissions with demographic features, length of stay, cost analysis, comorbidities, overall and gender-specific mortality and complications of transcutaneous Tricuspid replacement/repair [TTVR] vs. open surgical tricuspid valve replacement/repair [Open TVR].

**Methods:** Data was extrapolated from the NRD databases 2015-19. Of the 75,266,750 (unweighted) cases recorded in the 2015 – 2019 dataset, 429 had one or more of the percutaneous approach codes as per the ICD-10 data set, and 10077 had one or more of the open approach codes.

**Results:** Overall, the number of cases performed each year through open TVR was higher than TTVR, but there was an increased trend towards the TTVR every passing year. TTVR was performed more in females and advanced age groups than open TVR. The length of stay and cost was lower in the TTVR group than in open TVR. Patients undergoing TTVR had more underlying comorbidities like CHF, HTN, and uncomplicated DM. Overall mortality was 3.49 % in TTVR vs. 6.09% in open TVR. Gender-specific analysis demonstrated higher female mortality in the open TVR compared to TTVR (5.45% vs. 3.03 %). Male mortality was statistically insignificant between the two groups (6.8%% vs. 4.3%, p-value 0.15%). Patients with TTVR had lower rates of complications than open TVR, except for arrhythmias, which were higher in TTVR. Patients undergoing open TVR required more intracardiac support, such as IABP and Impella, than TTVR.

**Conclusion:** Transcatheter tricuspid valve replacement/repair is an emerging alternative to open surgical repair/replacement in patients with tricuspid valve diseases, especially tricuspid regurgitation. Despite having more underlying comorbidities, the TTVR group had lower in-hospital mortality, hospital cost, length of stay, and fewer complications than open TVR.

## Introduction

Recent data demonstrates that TV pathology especially tricuspid regurgitation (TR) is associated with poor long-term survival if left untreated **[1]**. Intervention on diseased Tricuspid valves, whether open or trans-catheter, has shown to improve the quality of life and severity of the valvular disease in these patients **[2]**. Tricuspid stenosis is uncommon and accounts only for 2.4% of tricuspid valve diseases **[3,4]** but Tricuspid regurgitation is very common and it affects > 1.6 million people in United States and > 70 million people worldwide **[5]**. The causes of primary TR include rheumatic heart disease, direct valvular injury during procedures, infective endocarditis, and connective tissue disorders **[6]**. Secondary TR is often functional from right ventricular dilatation secondary to left-sided heart diseases or diseases of the pulmonary system with normal tricuspid valve leaflet anatomy **[7]**. Tricuspid stenosis (TS) is rare and usually is due to congenital or acquired diseases affecting less than 1% of the general population in the developed world due to a decrease in the prevalence of rheumatic heart diseases **[8]**. Data from European Society Of Cardiology demonstrates that Surgical intervention is recommended for severe symptomatic TR and severe symptomatic TS either alone or at the time of surgery for left-sided valvular heart diseases **[9]**. Due to high surgical mortality associated with these valvular interventions, medical management was preferred in the past over surgery in the vast majority of these patients **[10]**. However, there has been a recent increase in trans-catheter tricuspid valve interventions for severe symptomatic tricuspid valvular diseases, especially in patients at higher risk with isolated tricuspid valve disease **[11,12]**. These interventions did show an improvement in functional status and reduction in severity of valvular disease and mortality **[13,14]**. Although, we have seen significant advances in trans-catheter treatments for mitral and aortic valves, transcatheter interventions for tricuspid valve are still in the developmental phase **[15]**. There is also limited data on head-to-head comparison of transcatheter vs. open tricuspid valve interventions. Therefore, we sought to use the Nation-wide readmission data base dataset to identify for the first time ever, the demographic features including morbidity and mortality analysis of trans-catheter vs. open surgical TV interventions.

## Materials and Methods

We used the NRD dataset from years 2015 – 2019 was used for this research project. NRD is a publicly available data sponsored by Agency for Healthcare Research and Quality. The database was developed for the HCUP [healthcare cost and utilization project] and it houses data on 35 million annual weighted discharges from around 28 States. Each patient is assigned a unique identifier code to trace readmissions within specific calendar year. Given the deidentified nature of the database, Institutional Review Board approval and Informed Consent were not required for this study. For each of the variables of interest, we calculated the Weighted Mean or Percentage, and Weighted Standard Error (SE), within each subgroup. Subgroups were cases with the Percutaneous Approach, cases with the Open Approach, or cases with neither. Z-Test Calculator was used to produce a p-value comparing the Percutaneous to Open Approach on the variables of interest. Of the 75,266,750 (unweighted) cases recorded in the 2015 – 2019 dataset, 429 had one or more of the percutaneous approach codes as per ICD-10 data set, and 10077 had one or more of the open approach codes.

## Results

### Baseline Characteristics of Patients

Data on demographics and baseline co-morbidities demonstrated significant differences between the two groups as shown in table **[Table 1]**. Patients undergoing open TVR were relatively younger as compared to TTVR group [mean age 40.8 vs 57.8 years, p-value <0.001] but there were more females in the TTVR group[66.04% vs 55.20%, p-value <0.001]. Data also revealed that patients in TTVR group were significantly sicker with more comorbidities as shown below. The percentage of CHF, uncomplicated HTN, OSA and uncomplicated DM respectively was 75.43%, 61.16%, 16.1% and 8.74% in the TTVR group as compared to 53.48%, 45.53%, 7.9% and 5.41% in the open TVR group**[Table 1]**. In contrast, complicated DM and complicated hypertension was higher in the open TVR group [0.83% and 17.38% vs 0% and 11.03%]. Patients in the TTVR group also had a trend towards higher prevalence of COPD but data was not statistically significant[19.9 vs 16.6, p-value 0.1089].

**Table 1:** Baseline Characteristics of patients in TTVR vs Open TVR.

| Variables | Percutaneous Approach | Open Approach | p-value |
| --- | --- | --- | --- |
| **Age** |  |  |  |
| **Mean year** | 57.8 (1.2458) | 40.8 (1.2752) | <0.001 |
| **Gender %** |  |  |  |
| **Females** | 66 (2.4172) | 55.2 (0.6283) | <0.001 |
| **Comorbidities %** |  |  |  |
| **CHF** | 75.4 (2.3292) | 53.4 (1.1088) | <0.001 |
| **DM - uncomplicated** | 8.74 (1.5020) | 5.41 (0.2946) | 0.029 |
| **DM - Complicated** | 0 (0) | 0.08 (0.0266) | 0.002 |
| **COPD** | 19.9 (1.9635) | 16.6 (0.6808) | 0.1089 |
| **HTN** | 61.1 (2.8370) | 45.5 (1.1936) | <0.001 |
| **HTN - complicated** | 11 (1.5900) | 17.3 (0.5095) | <0.001 |
| **HTN - Uncomplicated** | 50.5 (3.0331) | 28.9 (1.0479) | <0.001 |
| **OSA** | 16.1 (1.9607) | 7.96 (0.3721) | <0.001 |

- Standard error in parenthesis

### National Estimates for Percentage of All Readmissions

Of the 75,266,750 total cases, 429 underwent TTVR, and 10077 underwent open TVR. Although overall, the number of cases performed each year through open TVR was higher than TTVR, there was an increased trend towards the TTVR with every passing year (except for a small decrease in 2016) as shown in **[Table 2]**.

**Table 2:**
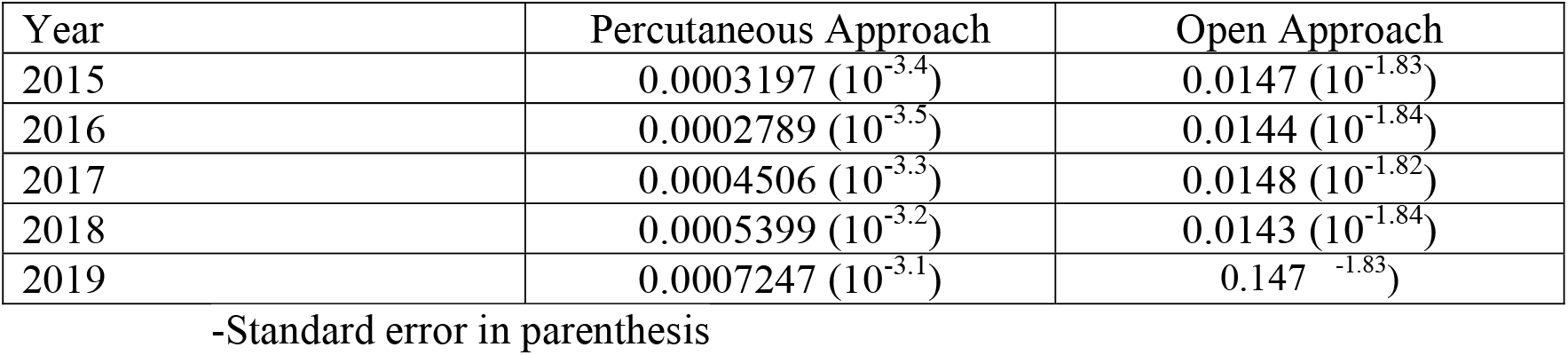
National Estimate for Percentage of all Readmissions for TTVR vs Open TVR.

| Year | Percutaneous Approach | Open Approach |
| --- | --- | --- |
| 2015 | 0.0003197 ($10^{-3.4}$) | 0.0147 ($10^{-1.83}$) |
| 2016 | 0.0002789 ($10^{-3.5}$) | 0.0144 ($10^{-1.84}$) |
| 2017 | 0.0004506 ($10^{-3.3}$) | 0.0148 ($10^{-1.82}$) |
| 2018 | 0.0005399 ($10^{-3.2}$) | 0.0143 ($10^{-1.84}$) |
| 2019 | 0.0007247 ($10^{-3.1}$) | 0.147 ($10^{-1.83}$) |

-Standard error in parenthesis

### In hospital outcomes

Among our patient population, the overall mortality was 3.49 % among the TTVR group vs. 6.09% in open TVR. Gender-specific analysis did demonstrate a higher female mortality in the open TVR as compared to TTVR group (5.45% vs 3.03 %, p-value 0.023). There was however, no statistical difference in mortality among males (6.8%% vs 4.3%, p-value 0.15)**[Table 3]**. Overall length of inpatient stay was also more than double in open TVR as compared to TTVR group (23.07 vs 9.8 days, p-value <0.001) **[Table 3]**. Cost analysis among the cases from the year 2019 (n=2471 open TVR, n=150 TTVR) demonstrated significantly higher mean costs in the open group vs TTVR. (mean 509107$ versus 308394) respectively (**Table 3**).

**Table 3:** In Hospital Outcomes of TTVR vs Open TVR.

| **Variables** |  | **Percutaneous Approach** | **Open Approach** | **p-value** |
| --- | --- | --- | --- | --- |
| **Mean LOS in days** |  | 9.85 (0.7845) | 23 (0.4566) | <0.001 |
| **Overall Mortality %** |  | 3.49 (0.8874) | 6.09 (0.3066) | 0.0057 |
|  | Female Mortality | 3.03 (1.0185) | 5.45 (0.3812) | 0.023 |
|  | Male Mortality | 4.39 (1.6394) | 6.82 (0.4528) | 0.1534 |
| **Cost in dollars** |  | 308394 | 509107 |  |

-Standard error in parenthesis

### Complications

Overall, patients in open TVR group had more complications as compared to TTVR group. The incidence of Cardiopulmonary Arrest was 2.27% in open TVR vs 1.05% in TTVR group. There was also an increased incidence of ARDS in the open TVR group (0.72% vs 0%, p-value <0.001). Interesting, no ARDS cases were reported in the TTVR group as shown in **[Table 4]**. ECMO requirements, however were almost similar in both groups(0.75% vs 0.70% in open TVR vs TTVR, p-value 0.907). In regards to major bleeds and blood loss anemia, data showed almost equal incidence in both groups [4.47% vs 4.2% and 1.2% vs 1.01% in open TVR vs TTVR, p-value 0.83 and 0.56 respectively].

**Table 4:** Complications of TTVR vs Open TVR.

| **Variables** | **Percutaneous Approach** | **Open Approach** | **p-value** |
| --- | --- | --- | --- |
| **Major bleed** | 4.2632 (1.0269) | 4.4781 (0.2580) | 0.839 |
| **Cardiac arrest** | 1.0505 (0.4580) | 2.7872 (0.1913) | <0.001 |
| **ARDS** | 0 (0) | 0.7294 (0.0939) | <0.001 |
| **Cardiac Tamponade** | 0.2127 (0.2113) | 1.9164 (0.1504) | <0.001 |
| **Pleural effusion** | 1.1908 (0.5212) | 8.3524 (0.3403) | <0.001 |
| **Arrhythmias** | 76.2786 (2.3614) | 62.2894 (1.1188) | <0.001 |
| **IABP** | 1.6158 (0.7511) | 4.7343 (0.3247) | <0.001 |
| **Impella** | 0.1907 (0.1912) | 0.5093 (0.0723) | 0.119 |
| **ECMO** | 0.7012 (0.4101) | 0.7506 (0.1093) | 0.907 |
| **Permanent pacemaker** | 4.1286 (0.9556) | 12.7029 (0.5321) | <0.001 |
| **Stroke** | 0.5738 (0.4165) | 1.4839 (0.1282) | 0.037 |
| **LVAD** | 0.4611 (0.3281) | 1.8043 (0.2020) | <0.001 |
| **Blood loss anemia** | 1.0130 (0.4428) | 1.2706 (0.1410) | 0.568 |
| **Infective endocarditis** | 3.57 (1.0050) | 25 | <0.001 |
| **Paravalvular leak** | 2.36 (0.8487) | 0.56 (0.0798) | 0.035 |

-Data are presented as the weighted percentage with standard error in parenthesis

A higher percentage of patients in the open TVR group required permanent pacemaker placement as compared to TTVR (12.7% vs 4.12%, p-value <0.001). Similarly, there was an increased incidence of pericardial effusion and cardiac tamponade in the open TVR population as compared to TTVR population (8.3% vs 1.1% and 1.9% vs 0.2%, p-value <0.001). Patients in the open TVR group also required more mechanical support as compared to TTVR group [IABP: 4.7% vs 1.6%, p-value < 0.001, LVAD: 1.8% vs 0.46%, p-value <0.001]. Data on requirement for Impella support was non-significant between the two groups but there was an increased trend noted in the Open group [0.5% vs 0.19%, p-value 0.119) as shown in **[Table 4]**. Patients in open TVR also had more Infective Endocarditis and a higher incidence of stroke as compared to TTVR group (25.04% vs 3.5 % and 1.4% vs 0.57, p-value <0.001 and 0.037 respectively). However, patients in the Trans-catheter group were found to have more post-op arrhythmias than in the open group (76.27% vs 62.2%, p-value <0.001). The incidence of para-valvular leak was also higher (2.3%) in the TTVR group as compared to 0.56% in open TVR (p-value 0.035).

## Discussion

Our analysis revealed that although the number of Trans-catheter interventions were significantly low as compared to open surgery but there was an increasing trend towards minimally invasive approach with each passing year (except for a small decrease in 2016). Patients in the trans-catheter group were also older and more sick which could explain the relative tendency to shift away from conventional surgery due to a higher risk of complications. Another important finding was that trans-catheter interventions were mostly performed in patients with isolated TR (11.17% vs 2.9%, p-value <0.001) as compared to conventional surgery. This is understandable as open surgery usually involves younger people with congenital heart disease with associated structural or valvular abnormalities whereas TR in old age is usually secondary to annular dilatation and may or may not be associated with other cardiac disease **[16, 17]**.

To our knowledge, this is the largest and first ever study comparing in-hospital outcomes and national readmission percentages of TTVR vs. open TVR in the American population. The major findings of our study are: An increased trend towards trans-catheter tricuspid interventions as compared to open surgery in years 2015-19, with a significantly lower overall and female mortality as compared to the open group. Trans-catheter approach was also deployed in relatively older patients as compared to open surgery, majorly for secondary TR likely secondary to annular dilation. It was also cheaper, with a shorter length of stay, hospitalization cost, and less complication rates.

The two common pathologies of the Tricuspid valve are regurgitation and stenosis. Tricuspid regurgitation is the more common pathology as compared to stenosis. The prevalence of TR increases with age, and it is also more prevalent in females **[18]**. Its management depends on the severity and etiology (primary vs. secondary) of the disease. Mild to moderate TR can be managed medically, while severe TR is managed with either surgical or trans-catheter intervention. As per the American College of Cardiology/American Heart Association joint Committee on clinical practice guidelines favours surgical repair over valve replacement if possible **[19]**. The choice of specific surgical technique depends on the stage of TR. Ring annuloplasty with prosthetic rings is usually performed at the mitral or aortic valve surgery in patients with mild to moderate TR with TA dilation and no significant tethering (coaptation height < 8 mm) **[20]**. But rigid undersize prosthetic rings may be used in patients with severe TA dilation(> 45mm) without significant leaflet tethering **[21]**. However, if valve replacement is indicated then bioprosthetic valves are preferred over mechanical valves due to low risk of thromboembolism. Controversy exists in literature regarding the appropriate timing of intervention for severe TR which is crucial to avoid irreversible damage to the right ventricle and worsening heart failure **[22]**. In the past, severe TR was usually medically managed with preload reduction including diuretics due to the high mortality associated with surgical intervention. There has been, however an increasing trend towards surgical repair of symptomatic severe TR, especially during surgical intervention for left-sided valvular heart diseases **[23]**. Studies have reported a poor prognosis in these patients if TR is left untreated during intervention for left-sided valvular heart diseases **[24]**. The European Society of Cardiology’s Valvular Heart Disease Guidelines recommend TV surgical intervention as a class 1C recommendation for symptomatic severe TS (especially during left-sided valve surgery), severe primary TR undergoing left-sided valve intervention, isolated severe primary TR without severe RV dysfunction and as a class 1B recommendation for severe secondary TR undergoing left-sided valve surgery **[25]**. Recent studies have shown that Trans-catheter tricuspid valve intervention can be used as an alternative option in select patients deemed surgically poor candidates **[7]**. A study performed on trans-catheter tricuspid valve interventions by Taramasso et al. showed that TTVR has low overall mortality and good functional outcomes with reasonable success rate in patients with severe TR **[26]**. Another study by Taramasso et al. showed that the all-cause mortality and 1-year rehospitalization were lower with TTVR as compared to medical management in patients with symptomatic severe TR **[27]**. Our analysis also demonstrated similar findings with lower mortality and morbidity outcomes with the trans-catheter approach.

We found TTVR being performed more frequently in females and older age populations as compared to open TVR because of the higher prevalence of TR and surgical inoperability in those groups**[18]**. There was also a higher prevalence of comorbidities in patients in the TTVR group compared to the open TVR group. The most common comorbidities were congestive heart failure (CHF), diabetes mellitus (DM), and hypertension (HTN). Especially, patients with LVEF < 40%, right ventricular dysfunction and or pulmonary hypertension carry higher a surgical risk and thus may benefit more from TTVR **[28]**. It also explains the higher prevalence of heart failure in patients undergoing TTVR in our study.

Prior studies have shown significant morbidity and mortality with surgical interventions for tricuspid valve diseases. Some retrospective studies have reported an in-hospital mortality of 10.9 % and 8.1 % with surgical isolated tricuspid valve replacement and repair, respectively **[29]**. This can be as high as ≥ 20-30 % in patients with pre-operative right ventricular dysfunction. Another study showed an in-hospital mortality of 8 % in patients undergoing tricuspid valve annuloplasty, while the mortality was 37 % in cases of reoperation (due to failure) **[30]**. The main prognostic factors contributing to mortality post-surgery were the presence of pre-operative right ventricular dysfunction, pre-operative organ dysfunction (for example, renal or liver dysfunction), underlying comorbidities, and reduced left ventricular function **[31]**. Late presentation for tricuspid valve surgery itself was a risk factor for higher mortality **[32]**. Our study showed an overall mortality of 3.49 % in TTVR vs. 6.09% in open TVR. Previous studies performed on transcatheter tricuspid interventions have reported varying degrees of mortality. One study reported an in-hospital mortality of 10 % **[33]** while another reported it to be around 13 % **[34]**. The mortality in transcatheter interventions depends on whether replacement or repair was done. A study has shown almost no mortality with the repair. The mortality with valve replacement was 5.7 % and 12.5 % with LUX and NAVIGATE valve systems, respectively **[35]**. Our study showed low mortality with TTVR despite more underlying comorbidities in that group.

Study by Zack et al. showed that higher hospitalization cost directly correlates with the length of stay, utilization of pacemakers, and in-hospital mortality **[10]**. In our study, the cost was higher for patients with open TVR as compared to TTVR, which can be explained by the higher length of stay, higher mortality, and higher incidence of permanent pacemaker placement in the open group. Similarly, the length of stay depends on the approach for TV interventions and pre-operative right ventricular function. Studies have shown an increased length of stay in patients with right ventricular dysfunction at the time of intervention **[36]**. A study performed by Fu et al. showed higher rates of cardiopulmonary bypass time, longer ICU stay, and longer ventilation time for patients with TV replacement as compared to repair, **[37]** which can also contribute to the longer length of stay. Our analysis showed that the length of stay in an open TVR was almost double that of TTVR [23.07 vs. 9.8]. A study performed by Buğan et al. found that the average length of stay was 10.7 days in patients with TTVR, which is similar to our study **[14]**.

Data shows that during cardiac surgery, around 0.2% to 6% of patients can develop post cardiotomy cardiogenic shock, this is characterized by tissue hypo-perfusion and end-organ damage despite adequate preload **[38]**. It is usually treated with vasopressors, ionotropic support, or in some cases, mechanical support such as an intra-aortic balloon pump insertion (IABP). Around 0.5 – 1% of these patients can also develop refractory post-cardiotomy cardiogenic shock not responsive to these measures, in which case, accelerated support such as ECMO is needed **[39,40]**. Patients with pre-operative right ventricular dysfunction are at higher risk of requiring intracardiac support, such as IABP and Impella, due to low cardiac output associated with right ventricular dysfunction **[41]**. In our study, patients who underwent open TVR were more likely to require an intra-aortic balloon pump (IABP) and Impella (although statistically insignificant) than TTVR. ECMO support was almost similar in both groups and statistically insignificant. Different studies have shown poor prognosis and higher short and long-term mortality in patients who required IABP or ECMO support after TAVR, **[42]** but there is limited such data on TTVR.

We also found high utilization of permanent pacemakers in patients with open TVR compared to TTVR. Several factors implicated in the requirement of a pacemaker during TV surgery include intra-operative hypothermia, duration of cardiopulmonary bypass, and proximity of the tricuspid annulus to the atrioventricular node **[43]**. The main reason for PPM placement is a complete heart block resulting from compression of the AV node by valve frame or ventricular anchors due to its proximity to the septal leaflet of the tricuspid valve **[44]**. Other predictors of PPM placement are infective endocarditis leading to AV block and baseline heart rhythm disturbance **[45]**. Previous studies have reported similar findings of permanent pacemaker requirement after valvular heart surgery. Two different studies on clinical outcomes of TV surgery showed that 21 % and 28 % of patients required permanent pacemakers **[46,47]**. Our study showed that 12 % of the patients required permanent pacemakers after the open TVR. A study on 3420 patients has reported that 14.1 % of patients underwent permanent pacemaker implantation after transcatheter aortic valve replacement (TAVR) **[48]**. Other studies have reported a 9 – 26 % prevalence of permanent pacemaker placement after TAVR **[49,50,51]**. But there is limited data on the need for permanent pacemakers after TTVR.

Pericardial effusion is a common complication after open cardiac surgery and can occur in up to 80% of patients **[52]**. The exact mechanism for the development of pericardial effusion is unknown, but the mechanical process of surgery and the inflammatory cytokines are thought to play an important role **[53]**. Anticoagulation also increases the risk of significant pericardial effusion and the development of cardiac tamponade **[54,55,56]**. Our study found that patients with open TVR have a higher incidence of pericardial effusions and cardiac tamponade than TTVR. This is understandable as open surgery by nature is more invasive and results in significant disruption of intra-cardiac environment and homeostasis as compared to the trans-catheter approach. A retrospective observational cohort study on 1460 patients showed that 16 % of patients undergoing heart valve surgery developed significant pericardial effusion requiring drainage **[57]**. A study on isolated tricuspid valve surgery reported that 8 % of patients developed cardiac tamponade after the surgery **[58]**. There is, however limited data available on the incidence of tamponade with transcatheter tricuspid valve interventions.

Patients with intra-cardiac valve replacement also have a significant risk of infective endocarditis (IE). This is usually due to the presence of resulting foreign material in the valve, subsequent paravalvular leaks, and damage to the native calcified valves from valve insertion and subclinical thrombosis **[59,60]**. Some 10 – 30 % of all infective endocarditis cases are caused by surgical prosthetic valve endocarditis. Numerous studies have shown a 5 – 50 % in-hospital mortality from infective endocarditis post valvular surgery **[61,62]**. Our analysis also showed a higher incidence of infective endocarditis in patients with open TVR than in the TTVR group. There is, although limited data in literature on the incidence of IE from trans-catheter tricuspid interventions. A recent review article on TAVRs which is the best studied among Trans-catheter interventions, showed that there was no difference in the incidence of prosthetic valve endocarditis between surgical and transcatheter aortic valve replacement **[63]**. Another study from a large multicenter registry showed the 1-year incidence of IE post-TAVR to be 0.50 % **[64]**. Further studies, however are needed in our case to elucidate any differences in IE between the trans-catheter and open groups if any.

Stroke is a rare but serious complication after interventions for valvular heart diseases, which increases morbidity and mortality in these patients. Due to the emerging nature of procedures, there is limited data on the risk of stroke with right-sided transcatheter valvular interventions. The available data on transcatheter tricuspid valve interventions have shown the incidence of the stroke to be close to 1 %, but the number of patients included in those studies was low **[26,65]**. Data from interventions for left-sided valvular heart diseases showed a relatively higher incidence of stroke. This could be due to a higher prevalence of atrial fibrillation in left-sided valvular heart diseases or a higher risk of atrial fibrillation from those interventions **[66]**. A multicentric German TAVI registry study showed the incidence of cerebrovascular events to be 3.2 %, with significantly higher in-hospital and 1-year mortality in patients developing cerebrovascular events **[67]**. The risk of stroke has been reported to be higher with surgical as compared to transcutaneous approaches in those patients **[49,68]**. Our study showed a higher incidence of stroke from open TVR than TTVR. However, more studies will be needed to accurately predict the risk of stroke in tricuspid valvular interventions.

## Conclusion

Transcatheter tricuspid valve replacement/repair is an emerging alternative to open surgical repair/replacement in patients with tricuspid valve diseases, especially high-risk populations with severe tricuspid regurgitation, to improve the quality of life. Our study’s analysis of a large pool of NRD data has shown promising trends towards lower morbidity and mortality and lower overall healthcare cost burden with TTVR compared to open TVR.

## Limitations

The administrative data and retrospective observational study design have their inherent limitations. As hospitalizations and not individual patients are represented in the data, there is a potential for overestimating the number of patients. The number of patients with TTVR in our study cohort was much smaller than open TVR, which could potentially have affected the significance of certain comparisons due to low power. However, the considerable sample size obtained from these large databases attenuates most of the limitations.

## Data Availability

The data that support the findings of this study are available from the corresponding author, [MK], upon reasonable request.

## References

1. Vassileva CM, Shabosky J, Boley T, Markwell S, Hazelrigg S. Tricuspid valve surgery: the past 10 years from the Nationwide Inpatient Sample (NIS) database. J Thorac Cardiovasc Surg. 2012;143(5):1043–1049. doi:10.1016/j.jtcvs.2011.07.004.

2. Nath J, Foster E, Heidenreich PA. Impact of tricuspid regurgitation on long-term survival. J Am Coll Cardiol. 2004;43(3):405–409. doi:10.1016/j.jacc.2003.09.036

3. Hauck AJ, Freeman DP, Ackermann DM, Danielson GK, Edwards WD. Surgical pathology of the tricuspid valve: a study of 363 cases spanning 25 years. Mayo Clin Proc. 1988;63(9):851–863. doi:10.1016/s0025-6196(12)62687-1

4. Golamari R, Shams P, Bhattacharya PT. Tricuspid Stenosis. In: StatPearls. Treasure Island (FL): StatPearls Publishing; January 4, 2023.

5. Demir OM, Regazzoli D, Mangieri A, et al. Transcatheter Tricuspid Valve Replacement: Principles and Design. Front Cardiovasc Med. 2018;5:129. Published 2018 Sep 19. doi:10.3389/fcvm.2018.00129.

6. Mulla S, Asuka E, Bora V, Siddiqui WJ. Tricuspid Regurgitation. In: StatPearls. Treasure Island (FL): StatPearls Publishing; November 17, 2022.

7. Agarwal S, Tuzcu EM, Rodriguez ER, Tan CD, Rodriguez LL, Kapadia SR. Interventional cardiology perspective of functional tricuspid regurgitation [published correction appears in Circ Cardiovasc Interv. 2010 Feb;3(1):e1. Tan, Carmela D [added]]. Circ Cardiovasc Interv. 2009;2(6):565–573. doi:10.1161/CIRCINTERVENTIONS.109.878983

8. Cevasco M, Shekar PS. Surgical management of tricuspid stenosis. Ann Cardiothorac Surg. 2017;6(3):275–282. doi:10.21037/acs.2017.05.14

9. Joint Task Force on the Management of Valvular Heart Disease of the European Society of Cardiology (ESC); European Association for Cardio-Thoracic Surgery (EACTS), Vahanian A, et al. Guidelines on the management of valvular heart disease (version 2012). Eur Heart J. 2012;33(19):2451–2496. doi:10.1093/eurheartj/ehs109

10. Zack CJ, Fender EA, Chandrashekar P, et al. National Trends and Outcomes in Isolated Tricuspid Valve Surgery. J Am Coll Cardiol. 2017;70(24):2953–2960. doi:10.1016/j.jacc.2017.10.039

11. Rodés-Cabau J, Hahn RT, Latib A, et al. Transcatheter Therapies for Treating Tricuspid Regurgitation. J Am Coll Cardiol. 2016;67(15):1829–1845. doi:10.1016/j.jacc.2016.01.063

12. Asmarats L, Puri R, Latib A, Navia JL, Rodés-Cabau J. Transcatheter Tricuspid Valve Interventions: Landscape, Challenges, and Future Directions. J Am Coll Cardiol. 2018;71(25):2935–2956. doi:10.1016/j.jacc.2018.04.031

13. Sorajja P, Whisenant B, Hamid N, et al. Transcatheter Repair for Patients with Tricuspid Regurgitation [published online ahead of print, 2023 Mar 4]. N Engl J Med. 2023;10.1056/NEJMoa2300525. doi:10.1056/NEJMoa2300525.

14. Buğan B, Çekirdekçi Eİ, Onar LÇ, Barçın C. Transcatheter Tricuspid Valve Replacement for Tricuspid Regurgitation: A Systematic Review and Meta-analysis. Anatol J Cardiol. 2022;26(7):505–519. doi:10.5152/AnatolJCardiol.2022.1440

15. Van Praet KM, Stamm C, Starck CT, et al. An overview of surgical treatment modalities and emerging transcatheter interventions in the management of tricuspid valve regurgitation. Expert Rev Cardiovasc Ther. 2018;16(2):75–89. doi:10.1080/14779072.2018.1421068

16. Singh JP, Evans JC, Levy D, et al. Prevalence and clinical determinants of mitral, tricuspid, and aortic regurgitation (the Framingham Heart Study) [published correction appears in Am J Cardiol 1999 Nov 1;84(9):1143]. Am J Cardiol. 1999;83(6):897–902. doi:10.1016/s0002-9149(98)01064-9.

17. Prihadi EA, Delgado V, Leon MB, Enriquez-Sarano M, Topilsky Y, Bax JJ. Morphologic Types of Tricuspid Regurgitation: Characteristics and Prognostic Implications. JACC Cardiovasc Imaging. 2019;12(3):491–499. doi:10.1016/j.jcmg.2018.09.027

18. Topilsky Y, Maltais S, Medina Inojosa J, et al. Burden of Tricuspid Regurgitation in Patients Diagnosed in the Community Setting. JACC Cardiovasc Imaging. 2019;12(3):433–442. doi:10.1016/j.jcmg.2018.06.014

19. Otto CM, Nishimura RA, Bonow RO, et al. 2020 ACC/AHA Guideline for the Management of Patients With Valvular Heart Disease: A Report of the American College of Cardiology/American Heart Association Joint Committee on Clinical Practice Guidelines [published correction appears in Circulation. 2021 Feb 2;143(5):e229]. Circulation. 2021;143(5):e72–e227. doi:10.1161/CIR.0000000000000923.

20. Tang GH, David TE, Singh SK, Maganti MD, Armstrong S, Borger MA. Tricuspid valve repair with an annuloplasty ring results in improved long-term outcomes. Circulation. 2006;114(1 Suppl):I577–I581. doi:10.1161/CIRCULATIONAHA.105.001263.

21. Ghoreishi M, Brown JM, Stauffer CE, et al. Undersized tricuspid annuloplasty rings optimally treat functional tricuspid regurgitation. Ann Thorac Surg. 2011;92(1):89–96. doi:10.1016/j.athoracsur.2011.03.024.

22. Dreyfus J, Ghalem N, Garbarz E, et al. Timing of Referral of Patients With Severe Isolated Tricuspid Valve Regurgitation to Surgeons (from a French Nationwide Database). Am J Cardiol. 2018;122(2):323–326. doi:10.1016/j.amjcard.2018.04.003

23. Baumgartner H, Falk V, Bax JJ, et al. 2017 ESC/EACTS Guidelines for the management of valvular heart disease. Eur Heart J. 2017;38(36):2739–2791. doi:10.1093/eurheartj/ehx391

24. Topilsky Y, Nkomo VT, Vatury O, et al. Clinical outcome of isolated tricuspid regurgitation. JACC Cardiovasc Imaging. 2014;7(12):1185–1194. doi:10.1016/j.jcmg.2014.07.018

25. Vahanian A, Beyersdorf F, Praz F, et al. 2021 ESC/EACTS Guidelines for the management of valvular heart disease [published correction appears in Eur J Cardiothorac Surg. 2022 Mar 24;61(4):964] [published correction appears in Eur J Cardiothorac Surg. 2022 Jun 15;62(1):]. Eur J Cardiothorac Surg. 2021;60(4):727–800. doi:10.1093/ejcts/ezab389

26. Taramasso M, Alessandrini H, Latib A, et al. Outcomes After Current Transcatheter Tricuspid Valve Intervention: Mid-Term Results From the International TriValve Registry. JACC Cardiovasc Interv. 2019;12(2):155–165. doi:10.1016/j.jcin.2018.10.022

27. Taramasso M, Benfari G, van der Bijl P, et al. Transcatheter Versus Medical Treatment of Patients With Symptomatic Severe Tricuspid Regurgitation. J Am Coll Cardiol. 2019;74(24):2998–3008. doi:10.1016/j.jacc.2019.09.028

28. Henning RJ. Tricuspid valve regurgitation: current diagnosis and treatment. Am J Cardiovasc Dis. 2022;12(1):1-18. Published 2022 Feb 15.

29. Alqahtani F, Berzingi CO, Aljohani S, Hijazi M, Al-Hallak A, Alkhouli M. Contemporary Trends in the Use and Outcomes of Surgical Treatment of Tricuspid Regurgitation. J Am Heart Assoc. 2017;6(12):e007597. Published 2017 Dec 22. doi:10.1161/JAHA.117.007597.

30. McCarthy PM, Bhudia SK, Rajeswaran J, et al. Tricuspid valve repair: durability and risk factors for failure. J Thorac Cardiovasc Surg. 2004;127(3):674–685. doi:10.1016/j.jtcvs.2003.11.019

31. Taramasso M, Pozzoli A, Guidotti A, et al. Percutaneous tricuspid valve therapies: the new frontier. Eur Heart J. 2017;38(9):639–647. doi:10.1093/eurheartj/ehv766.

32. Kim JB, Jung SH, Choo SJ, Chung CH, Lee JW. Clinical and echocardiographic outcomes after surgery for severe isolated tricuspid regurgitation. J Thorac Cardiovasc Surg. 2013;146(2):278–284. doi:10.1016/j.jtcvs.2012.04.019

33. Hahn RT, Kodali S, Fam N, et al. Early Multinational Experience of Transcatheter Tricuspid Valve Replacement for Treating Severe Tricuspid Regurgitation. JACC Cardiovasc Interv. 2020;13(21):2482–2493. doi:10.1016/j.jcin.2020.07.008

34. Lu FL, An Z, Ma Y, et al. Transcatheter tricuspid valve replacement in patients with severe tricuspid regurgitation. Heart. 2021;107(20):1664–1670. doi:10.1136/heartjnl-2020-318199.

35. Rahgozar K, Ho E, Goldberg Y, Chau M, Latib A. Transcatheter tricuspid valve repair and replacement: a landscape review of current techniques and devices for the treatment of tricuspid valve regurgitation. Expert Rev Cardiovasc Ther. 2021;19(5):399–411. doi:10.1080/14779072.2021.1915133

36. Hahn RT, George I, Kodali SK, et al. Early Single-Site Experience With Transcatheter Tricuspid Valve Replacement. JACC Cardiovasc Imaging. 2019;12(3):416–429. doi:10.1016/j.jcmg.2018.08.034

37. Fu J, Chen Q, Xu D, Zhao F, Liu Z, Jiang N. Zhonghua Yi Xue Za Zhi. 2015;95(18):1396–1400.

38. Rao V, Ivanov J, Weisel RD, Ikonomidis JS, Christakis GT, David TE. Predictors of low cardiac output syndrome after coronary artery bypass. J Thorac Cardiovasc Surg. 1996;112:38–51.

39. Sidebotham D, McGeorge A, McGuinness S, Edwards M, Willcox T, Beca J. Extracorporeal membrane oxygenation for treating severe cardiac and respiratory disease in adults: part 1doverview of extracorporeal membrane oxygenation. J Cardiothorac Vasc Anesth 2009;23:886

40. Marasco SF, Lukas G, McDonald M, McMillan J, Ihle B. Review of ECMO (extra corporeal membrane oxygenation) support in critically ill adult patients. Heart Lung Circ 2008;17(Suppl. 4):S41.

41. Cheng Y, Mo S, Wang K, et al. Mid-Term Outcome after Tricuspid Valve Replacement. Braz J Cardiovasc Surg. 2020;35(5):644–653. Published 2020 Oct 1. doi:10.21470/1678-9741-2019-0215

42. Shreenivas SS, Lilly SM, Szeto WY, et al. Cardiopulmonary bypass and intra-aortic balloon pump use is associated with higher short and long term mortality after transcatheter aortic valve replacement: a PARTNER trial substudy. Catheter Cardiovasc Interv. 2015;86(2):316–322. doi:10.1002/ccd.25776

43. Jouan J, Mele A, Florens E, et al. Conduction disorders after tricuspid annuloplasty with mitral valve surgery: Implications for earlier tricuspid intervention. J Thorac Cardiovasc Surg. 2016;151(1):99–103. doi:10.1016/j.jtcvs.2015.09.063

44. Fam NP, von Bardeleben RS, Hensey M, et al. Transfemoral Transcatheter Tricuspid Valve Replacement With the EVOQUE System: A Multicenter, Observational, First-in-Human Experience. JACC Cardiovasc Interv. 2021;14(5):501–511. doi:10.1016/j.jcin.2020.11.045

45. Herrmann FEM, Graf H, Wellmann P, Sadoni S, Hagl C, Juchem G. Etiology of tricuspid valve disease is a predictor of bradyarrhythmia after tricuspid valve surgery. J Cardiovasc Electrophysiol. 2019;30(7):1108–1116. doi:10.1111/jce.13937

46. Jokinen JJ, Turpeinen AK, Pitkänen O, Hippeläinen MJ, Hartikainen JE. Pacemaker therapy after tricuspid valve operations: implications on mortality, morbidity, and quality of life. Ann Thorac Surg. 2009;87(6):1806–1814. doi:10.1016/j.athoracsur.2009.03.048.

47. Do QB, Pellerin M, Carrier M, et al. Clinical outcome after isolated tricuspid valve replacement: 20-year experience. Can J Cardiol. 2000;16(4):489–493.

48. Rück A, Saleh N, Glaser N. Outcomes Following Permanent Pacemaker Implantation After Transcatheter Aortic Valve Replacement: SWEDEHEART Observational Study. JACC Cardiovasc Interv. 2021;14(19):2173–2181. doi:10.1016/j.jcin.2021.07.043

49. Popma JJ, Deeb GM, Yakubov SJ, et al. Transcatheter Aortic-Valve Replacement with a Self-Expanding Valve in Low-Risk Patients. N Engl J Med. 2019;380(18):1706–1715. doi:10.1056/NEJMoa1816885.

50. Leon MB, Smith CR, Mack MJ, et al. Transcatheter or Surgical Aortic-Valve Replacement in Intermediate-Risk Patients. N Engl J Med. 2016;374(17):1609–1620. doi:10.1056/NEJMoa1514616.

51. Bekeredjian R, Szabo G, Balaban ü, et al. Patients at low surgical risk as defined by the Society of Thoracic Surgeons Score undergoing isolated interventional or surgical aortic valve implantation: in-hospital data and 1-year results from the German Aortic Valve Registry (GARY). Eur Heart J. 2019;40(17):1323–1330. doi:10.1093/eurheartj/ehy699.

52. Kuralay E, Ozal E, Demirkilic U, et al: Effect of posterior pericardiotomy on postoperative supraventricular arrhythmias and late pericardial effusion (posterior pericardiotomy). J Thorac Cardiovasc Surg 1999;118:492–495.

53. Karatolios K, Moosdorf R, Maisch B, Pankuweit S. Cytokines in pericardial effusion of patients with inflammatory pericardial disease. Mediators Inflamm. 2012;2012:382082. doi:10.1155/2012/382082.

54. Alkhulaifi AM, Speechly-Dick ME, Swanton RH, et al: The incidence of significant pericardial effusion and tamponade following major aortic surgery. J Cardiovasc Surg 1996;37:385–389.

55. Malouf JF, Alam S, Gharzeddine W, et al: The role of anticoagulation in the development of pericardial effusion and late tamponade after cardiac surgery. Eur Heart J 1993;14:1451–1457

56. Chidambaram M, Akhtar MJ, al-Nozha M, et al: Relationship of atrial fibrillation to significant pericardial effusion in valve-replacement patients. Thorac Cardiovasc Surg 1992;40:70–73.

57. Borregaard B, Sibilitz KL, Weiss MG, et al. Occurrence and predictors of pericardial effusion requiring invasive treatment following heart valve surgery. Open Heart. 2022;9(1):e001880. doi:10.1136/openhrt-2021-001880

58. Dreyfus J, Flagiello M, Bazire B, et al. Isolated tricuspid valve surgery: impact of aetiology and clinical presentation on outcomes. Eur Heart J. 2020;41(45):4304–4317. doi:10.1093/eurheartj/ehaa643

59. Makkar RR, Fontana GP, Jilaihawi H, et al. Transcatheter aortic-valve replacement for inoperable severe aortic stenosis [published correction appears in N Engl J Med. 2012 Aug 30;367(9):881]. N Engl J Med. 2012;366(18):1696–1704. doi:10.1056/NEJMoa1202277

60. Chakravarty T, Søndergaard L, Friedman J, et al. Subclinical leaflet thrombosis in surgical and transcatheter bioprosthetic aortic valves: an observational study. Lancet. 2017;389(10087):2383–2392. doi:10.1016/S0140-6736(17)30757-2

61. Abegaz TM, Bhagavathula AS, Gebreyohannes EA, Mekonnen AB, Abebe TB. Short- and long-term outcomes in infective endocarditis patients: a systematic review and meta-analysis [published correction appears in BMC Cardiovasc Disord. 2018 Jan 12;18(1):5]. BMC Cardiovasc Disord. 2017;17(1):291. Published 2017 Dec 12. doi:10.1186/s12872-017-0729-5

62. Habib G, Erba PA, Iung B, et al. Clinical presentation, aetiology and outcome of infective endocarditis. Results of the ESC-EORP EURO-ENDO (European infective endocarditis) registry: a prospective cohort study [published correction appears in Eur Heart J. 2020 Jun 7;41(22):2091]. Eur Heart J. 2019;40(39):3222–3232. doi:10.1093/eurheartj/ehz620.

63. Alexis SL, Malik AH, George I, et al. Infective Endocarditis After Surgical and Transcatheter Aortic Valve Replacement: A State of the Art Review. J Am Heart Assoc. 2020;9(16):e017347. doi:10.1161/JAHA.120.017347

64. Amat-Santos IJ, Messika-Zeitoun D, Eltchaninoff H, et al. Infective endocarditis after transcatheter aortic valve implantation: results from a large multicenter registry [published correction appears in Circulation. 2020 Jun 2;141(22):e879]. Circulation. 2015;131(18):1566–1574. doi:10.1161/CIRCULATIONAHA.114.014089

65. Wu Z, Zhu W, Kaisaier W, et al. Periprocedural, short-term, and long-term outcomes following transcatheter tricuspid valve repair: a systemic review and meta-analysis. Ther Adv Chronic Dis. 2023;14:20406223231158607. Published 2023 Mar 4. doi:10.1177/20406223231158607

66. Bando K, Kobayashi J, Hirata M, et al. Early and late stroke after mitral valve replacement with a mechanical prosthesis: risk factor analysis of a 24-year experience. J Thorac Cardiovasc Surg. 2003;126(2):358–364. doi:10.1016/s0022-5223(03)00550-6

67. Werner N, Zeymer U, Schneider S, et al. Incidence and Clinical Impact of Stroke Complicating Transcatheter Aortic Valve Implantation: Results From the German TAVI Registry. Catheter Cardiovasc Interv. 2016;88(4):644–653. doi:10.1002/ccd.26612

68. Mack MJ, Leon MB, Thourani VH, et al. Transcatheter Aortic-Valve Replacement with a Balloon-Expandable Valve in Low-Risk Patients. N Engl J Med. 2019;380(18):1695–1705. doi:10.1056/NEJMoa1814052

